# Community-Based Accompaniment for Adolescents Transitioning to Adult HIV Care in Urban Peru: a Pilot Study

**DOI:** 10.1101/2021.08.25.21261815

**Authors:** Valentina Vargas, Milagros Wong, Carly A. Rodriguez, Hugo Sanchez, Jerome Galea, Alicia Ramos, Liz Senador, Lenka Kolevic, Eduardo Matos, Eduardo Sanchez, Renato A. Errea, Karen Ramos, Catherine Beckhorn, Andrew Lindeborg, Carlos Benites, Leonid Lecca, Sonya Shin, Molly F. Franke

## Abstract

**Introduction:** Adolescents living with HIV (ALWH) face an elevated risk of poor health outcomes when transitioning into adult-oriented care; however, evidence-based interventions to support ALWH during this high-risk period are lacking, especially in Latin America. We conducted a pilot study of a community-based intervention designed to improve outcomes among ALWH transitioning to adult HIV care in Lima, Peru.

**Methods:** From October 2019 to January 2020, we enrolled adolescents transitioning to adult HIV care, either due to a recent diagnosis or having aged out of their pediatric clinic. ALWH previously lost from care during the transition process were also invited to participate. The nine-month intervention consisted of (1) logistical, adherence and social support delivered by entry-level health workers who accompanied adolescents during their transition to adult HIV care and (2) group sessions to improve health-related knowledge, skills and social support. We assessed intervention feasibility and effectiveness in improving medication adherence, psycho-social outcomes, and transition readiness after 6, 9, and 12 months.

**Results:** We enrolled 30 ALWH, ages 15-21 years; 11 were recently diagnosed and 19 had been living with HIV since infancy or early childhood. Participants included ten men who have sex with men, four young pregnant women and six adolescents who were previously lost during the transition to adult HIV care. Intervention participation was high with 90% of ALWH attending at least one in-person social support session and all attending at least one live online session. No ALWH withdrew from the intervention, study, or antiretroviral therapy. In transition readiness, we observed within-person improvements related to personal health (+1.9 points, p<0.001), healthcare usage (+2.4 points, p<0.001), knowledge (+3.3 points, p=0.001), and behavior (+3 points, p=0.003) at the end of the intervention, relative to baseline. We also observed strong evidence of improvements in medication adherence, social support, self-efficacy, and perceived stress, which were generally sustained three months after intervention cessation.

**Conclusion:** We identified a community-based intervention that is feasible and potentially effective for bridging the transition to adult HIV care among a diverse group of ALWH in Peru. A larger-scale effectiveness evaluation, including biological endpoints, is warranted.

## Introduction

Expanded access to antiretroviral therapy (ART) has led to major improvements in the survival of children who acquire HIV early in life. This means that providers and programs are faced with the challenge of caring for a group of patients living with HIV that is also dealing with the many developmental, emotional and behavioral issues of adolescence. These include the desire for peer acceptance, vulnerability to mental health and substance use disorders, and the potential consequences of early sexual debut (e.g., pregnancy, HIV transmission, other sexually-transmitted infections) [1–6]. An increased risk of early life adversity and trauma among adolescents living with HIV (ALWH) may make them especially susceptible to mental health morbidity [7,8]. And, ALWH may lack the life skills, coping strategies and support needed to effectively manage common HIV-related challenges, such as diagnosis disclosure, HIV-related stigma and social isolation, neurocognitive delays, and navigating romantic relationships [1–3,8–11]. In spite of their unique needs, ALWH have rarely been targeted for intervention studies to improve health outcomes [12– 14]. The absence of tailored interventions for adolescents may in part explain their disproportionately high rates of attrition to care relative to other age groups [15].

The transition from pediatric to adult care is an exceptionally precarious time in adolescent HIV care, marked by reduced clinic attendance, retention in care, HIV viral load suppression and CD4 cell counts [16,17]. This transition, which often occurs in late adolescence, can be accompanied by logistical and emotional challenges, as adolescents find themselves newly independent with different medical providers in an unfamiliar and often less supportive environment [18,19]. Although several reviews suggest good transition outcomes may be achieved with individualized transition plans that address the multifaceted needs of adolescence [8,19–22], there are few rigorous studies testing promising interventions [23,24] and a notable lack of evidence on which interventions might be most effective in supporting ALWH through this period.

Alongside an urgent need to identify effective transition intervention models is a growing recognition of the need for interventions that are tailored to local context and to specific subgroups. A recent review on care transition studies for ALWH identified an absence of studies from Latin America and the Caribbean, where the experiences of ALWH likely differ from those in high-income or high prevalence settings [24]. There is also a stark need for transition studies among adolescents with behaviorally-acquired HIV[8], including young men who have sex with men (MSM) and transgender women (TGW), as their transition experiences may differ from those of perinatally-infected adolescents [25]. As a first step toward addressing these research gaps, we conducted a pilot study to examine whether an intervention for ALWH, rooted in community-based support, could be a feasible approach for bridging the transition to adult HIV care among a diverse group of adolescents in an urban Latin American setting.

## Methods

### Study setting

In Peru, there are an estimated 87,000 people living with HIV, with the highest concentration in the capital, Lima [26,27]. While perinatal infection constitutes the majority of HIV infections in younger adolescents, most new infections in Peru occur among MSM and TGW, reflecting Peru’s concentrated epidemic [28]. Pregnancy during adolescence is common with 17% of 20- to 24-year-old women giving birth before age 18 [29]. ART is available free-of-charge and is largely centralized with most patients receiving treatment through public sector hospitals.

For adolescents that acquired HIV in early childhood HIV, the transition from a pediatric to an adult HIV clinic often occurs at age 18, though it may occur as young as 15 for adolescents transitioning within the same health facility. Older, newly-diagnosed adolescents may transition directly into adult HIV care, bypassing the pediatric clinic. If an adolescent in pediatric care becomes pregnant, she immediately transitions to an adult HIV clinic, regardless of age. The existing standard of care for adolescents transitioning to adult care includes a public health insurance referral and a series of consults with a nurse, infectious disease specialist, psychologist, and social worker prior to transition. The patient is given a report from each provider team, which the ALWH is responsible for bringing to their first adult clinic visit.

### Participants

We enrolled ALWH aged 15 to 21 years who were on or eligible for ART, enrolled in HIV care at a participating public sector clinic, and scheduled to transition to adult HIV care, either due to a recent HIV diagnosis or because they had aged out of their pediatric clinic. ALWH who were lost from adult HIV care at the time of transition but interested in re-engaging in treatment were also eligible to participate. Adolescents were excluded if they lived outside of metropolitan Lima. Enrollment took place from October 2019 to January 2020 at three public sector hospitals (one pediatric facility, and two facilities with distinct on-site pediatric and adult clinics). Ministry of Health (MoH) clinical collaborators consecutively referred eligible participants to study staff. A Youth Advisory Board (YAB), comprising a diverse group of young people living with HIV provided feedback on all aspects of the study. Adolescents who had previously withdrawn from care were referred by their clinicians or YAB members and contacted by study staff.

### Study Intervention

The intervention was nicknamed “PASEO” (meaning ‘crossing’ or ‘passage’ in Spanish) after its core elements: peer engagement, accompaniment, support and education. In short, PASEO aimed to provide in-person accompaniment to ALWH throughout the transition process through the provision of individualized support, including adherence support, to address barriers to care and treatment (e.g., transport costs, inaccessibility of services, lack of social support) by providing medical, material, and social support delivered by compensated entry-level or lay health workers who liaise with formal health services (Figure 1).[30]

**Figure 1.**
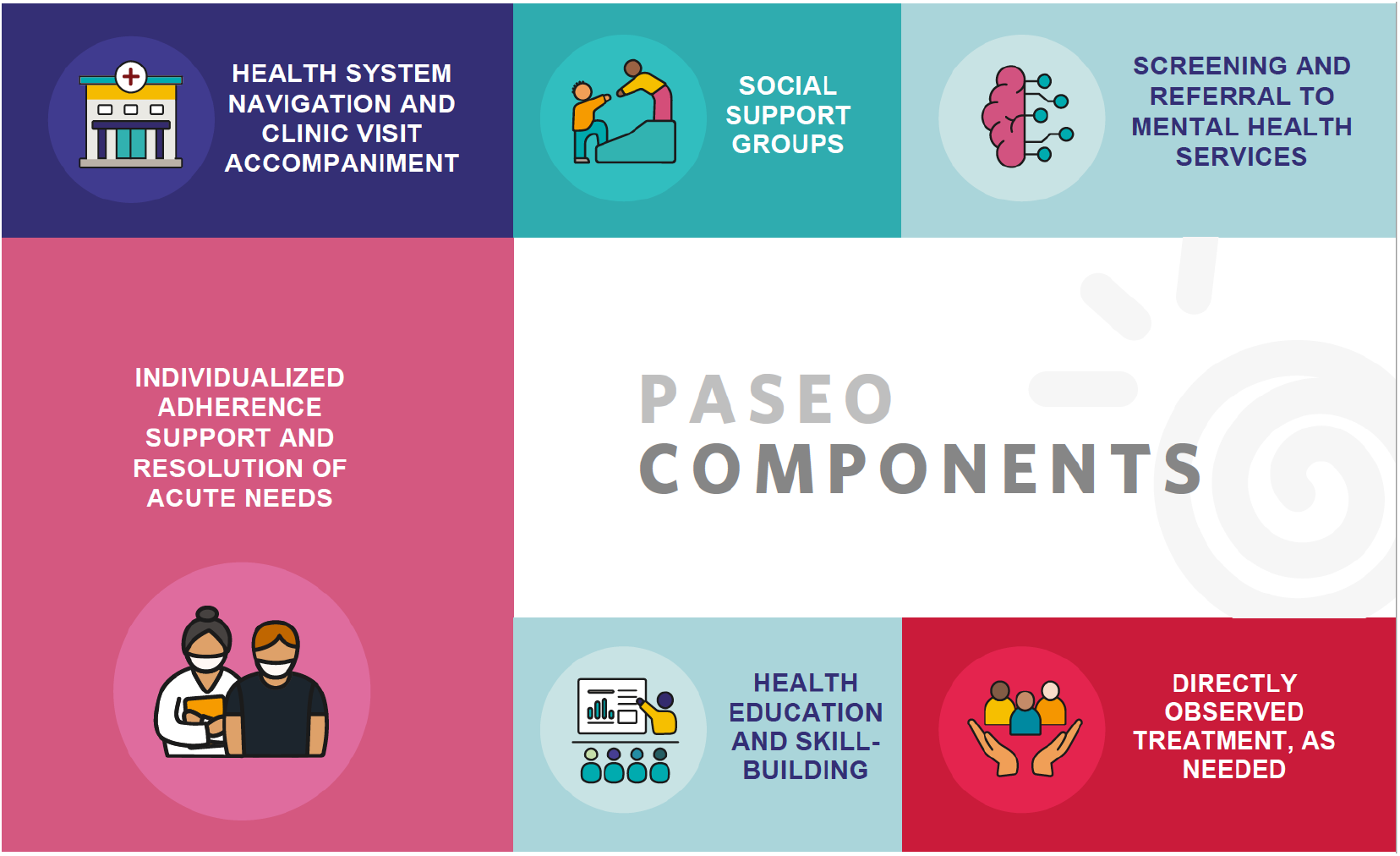
Key components of the PASEO intervention.

PASEO also aimed to foster health-related skills, knowledge, and psychosocial well-being needed to thrive in the adult clinic and more broadly. Intervention activities were delivered in two phases, an intensive phase and a taper phase (Figure 2). The intensive phase began just prior to transition (defined as the last date of care at the pediatric clinic or, for adolescents initiating treatment directly in an adult clinic, the ART initiation date) and ended after six months. A three-month taper phase, during which the intensity of study activities was reduced, followed the intensive phase. Below we describe each intervention component in detail (Figure 1), as well as adaptations made to accommodate stay-at-home orders invoked on March 15, 2020, due to the SARS-CoV-2 pandemic. Participants had received between 1.4 and 5.3 months of in-person intervention (median: 3.1 [IQR: 2.2, 4.2]) when in-person activities were suspended and were provided phone credit to enable participation in virtual programming.

**Figure 2.**
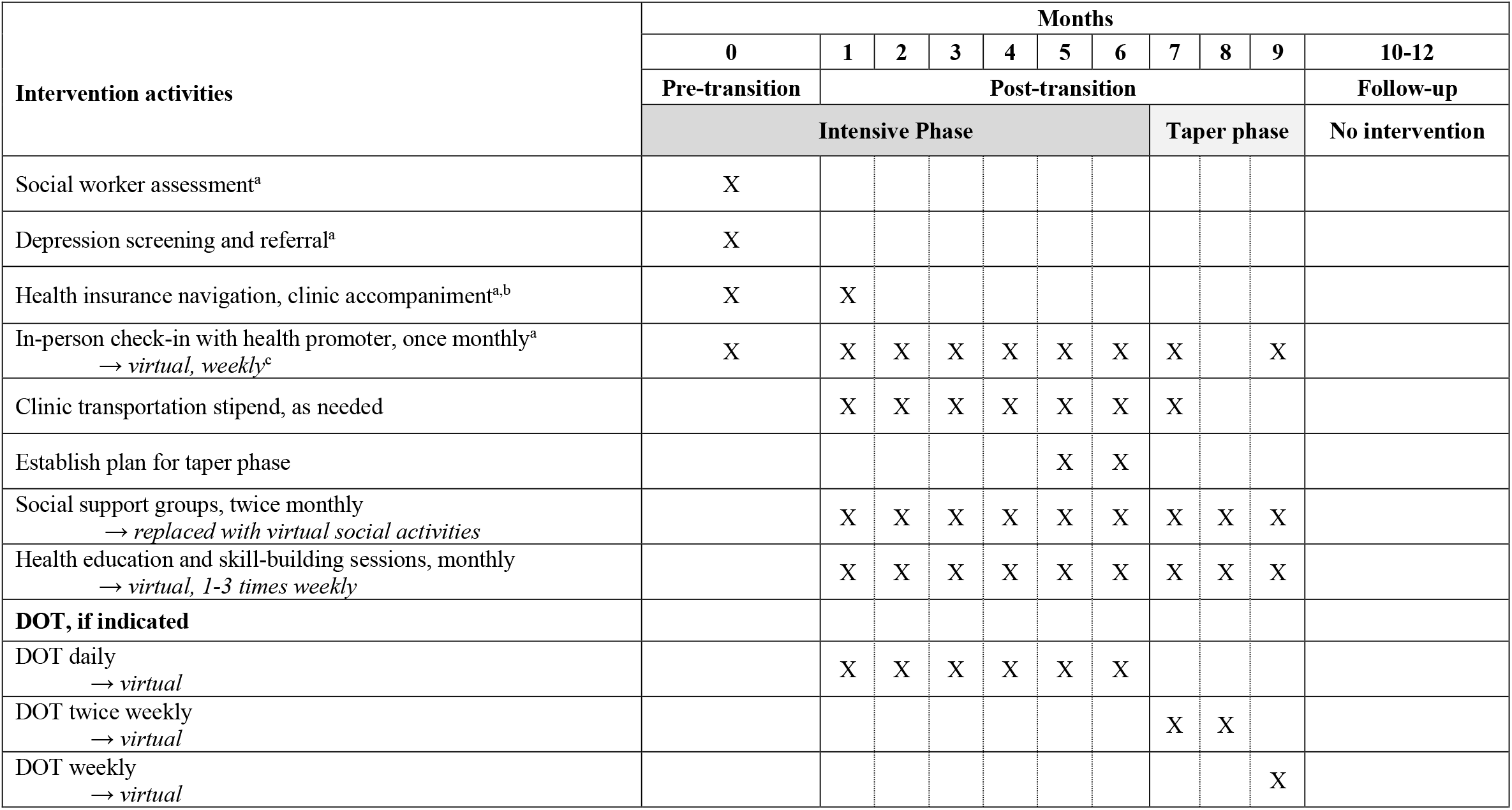
Overview of PASEO intervention implementation. DOT, Directly Observed Treatment a. Occurred more frequently, as needed b. Accompaniment to clinic visits typically occurred through months two or three, with longer durations for those previously lost from care, experienced adverse reactions or side effects of treatment, or had concomitant health issues. Clinic visit accompaniment occurred for shorter durations if the adolescent received this support from a caregiver or family member. c. Italicized text represents modifications made in response to the SARS-CoV-2 pandemic

### Health system navigation and clinic accompaniment

To facilitate the transition to care in the adult clinic, trained health promoters (entry-level health workers with a technical degree) employed by the nongovernmental organization Socios En Salud (SES) accompanied adolescents to their first appointments; facilitated enrollment in public health insurance and completion of other administrative requisites (e.g., obtaining a foreign identification card for migrants; transferring to a different health facility); helped troubleshoot new logistical and/or social challenges (e.g., housing instability); and served as a liaison between the patient and pediatric and adult clinical providers. Clinic visit transportation was provided based on economic need, as determined by the study social worker. All but one participant had attended at least one accompanied in-person clinic visit at the time SARS-CoV-2 stay-at-home orders were invoked. The exception was an individual who was scheduled to re-initiate care in mid-March 2020. While hospital guidelines precluded in-person accompaniment for this participant, intervention staff coordinated with HIV program staff to ensure that adolescent would be seen in the clinic. Study staff coordinated private telehealth appointments for adolescents requiring clinical consultation during stay-at-home orders.

### Routine check-in visits

Health promoters visited the participant’s home or another mutually-agreed upon location at least monthly, to review ART adherence, identify barriers to care and adherence, remind patients of upcoming medical encounters, screen and follow-up for clinical and social problems and offer social support. The frequency of check-ins was individualized and commensurate with the level of support needed. Visits tended to occur more frequently for participants lacking family support, re-initiating ART or with a history of unstable adherence.

### Directly Observed Treatment (DOT)

Home-based DOT was offered to adolescents with early childhood infection identified as at risk of non-adherence (based on pediatric health provider assessment or a detectable viral load at one of two previous measurements). ALWH newly initiating ART chose whether they wished to receive DOT. Eligible ALWH who opted into DOT were assigned to a MoH community health worker (CHW) from the same neighborhood to observe ART ingestion daily, ensure treatment was taken as prescribed and offer social support. Following invocation of stay-at-home orders, DOT was conducted virtually by SES health promotors via telephone, video or SMS, depending on participant preference. Some participants elected to participate in group video DOT, creating an additional opportunity to build social support.

### Social support groups, health education sessions and skill-building workshops

Social support groups took place twice monthly (10 adolescents per group). The two-hour sessions were facilitated by a Peruvian study psychologist and one peer youth living with HIV. Based on timing of enrollment, between two and five in-person sessions were offered to each adolescent prior to stay-at-home orders. Conducting these sessions online was largely unsuccessful: many adolescents felt they lacked privacy at home to discuss personal topics [31], and others could not use video due to unstable internet. To build and sustain social connectedness, staff implemented online social activities including an HIV-related meme competition, a TikTok video competition, and a virtual talent show.

Health education sessions and skill-building workshops were conducted on topics such as HIV, substance use, sexual health, self-esteem, nutrition, and gender and sexual identity. In-person sessions took place at least once monthly (coinciding with social support group sessions), while virtual sessions took place one to three times weekly (total of 41). For virtual sessions, participants could join live, allowing interaction with the speaker and other participants via chat, or view a recording.

### Mental health screening, referral and support

Participants were evaluated for depressive symptoms using the Patient Health Questionnaire-9 (PHQ-9), a tool previously validated in Spanish [32], and widely used in Peru [33]. Adolescents screening positive for mild or greater depressive symptoms (PHQ-9 ≥ 10) or with suicidal ideation were evaluated by a SES’ mental health program and offered Psychological First Aid [34] and referrals as needed. Participants with severe depressive symptoms (PHQ-9 ≥ 20) or suicidal ideation were offered linkage to free, specialized mental health services provided by the Peruvian Ministry of Health and delivered at nearby community mental health centers [35].

## Data Collection

Data collection took place from October 2019 to January 2021 and included indicators of intervention feasibility and effectiveness, and social, clinical, and demographic factors that could challenge transition.

Feasibility indicators aligned with selected dimensions of acceptability and demand [36]. Acceptability focuses on the extent to which a program is judged as suitable or attractive. We assessed acceptability through study refusal rates (i.e., the proportion of potentially eligible individuals for the study who chose not to participate) and intervention retention rates (i.e., the proportion of individuals who were retained in the study). The demand domain describes the extent to which a new intervention is likely to be used. We assessed demand through intervention retention rates and group attendance for study activities. Group attendance was assessed by the proportion of individuals who attended activities, the number who attended at least one session, the median number of groups attended by participants who attended at least one session, median number of participants per session, and the number of engagements (i.e., typed comments) during virtual sessions.

Preliminary evidence of effectiveness is an impact indicator examining whether a new intervention shows promise of success with the intended population. We generated preliminary evidence of effectiveness by calculating within-person changes in self-reported adherence, perceived social and instrumental support, self-efficacy and stress, and transition readiness at 6, 9, and 12-months, relative to baseline. Follow-up measurements corresponded to the end of the intensive phase, the end of the taper phase, and three months after intervention, respectively.

### Baseline clinical data

including CD4 cell count and viral load, were recorded from clinical charts. Follow-up measurements were rarely available due to testing disruptions resulting from the SARS-CoV-2 pandemic.

### Instruments

Participants self-administered surveys in Spanish on a tablet device. We assessed self-reported adherence during the prior 30-day period with three questions: “How many days did you miss at least one dose of any of your ART medications?”; “How often did you take your ART medications correctly?” (5-point scale with 5 representing “always” and 1 representing “never”); “How well would you say you took your HIV medications, as directed by your doctor?” (6-point scale with 6 representing “excellent” and 1 representing “very bad”). We assessed perceived instrumental support, emotional support, perceived stress, and self-efficacy using the Spanish fixed-form version of the NIH Toolbox Emotion Battery (version 2.0) for ages 18-85. All have a 5-point ordinal scale ranging from “never” to “always”, with higher scores indicating higher levels [37,38]. We evaluated transition readiness with two scales. “Am I On TRAC?” [39] consists of knowledge and behavior indices, which we adapted to the local context (see Appendix). The knowledge scale assesses the respondent’s health condition and general medical self-care. The behavior scale measures the frequency with which respondents engage in individual health-related behaviors related to the developmental model of transition. We also used the Got Transition? checklist (version 2.0), which assesses personal health knowledge and use of medical services [40]. The questionnaire has a 3-point scale ranging from “not my responsibility” to “yes, I know this.” For both measures, we report the summed score of responses for each subscale; higher scores indicate greater knowledge, engagement or use.

### Statistical Analysis

Feasibility indicators were reported as descriptive statistics. To examine changes in outcomes during the different phases of intervention implementation, we calculated within-person change from baseline to each of the three follow-up points and tested whether this quantity differed from zero, using paired t-tests or Wilcoxon signed rank tests, as appropriate. For the NIH toolbox measurements, we report raw scores instead of the t-values validated in a U.S. population. To understand whether potential effectiveness differed among subgroups, we stratified analyses by early childhood versus recent diagnosis and whether the adolescent had been lost from care or had a history of chronic non-adherence. Data were analyzed using SAS version 9.4 (Cary, NC).

### Compliance of Ethical Standards

The authors declare no conflicts of interest. Ethical approval was granted in Peru by the Institutional Research Ethics Committees of the Instituto Nacional de Salud del Niño, Hospital Nacional Arzobispo Loayza, and Hospital Nacional Hipólito Unanue. In addition, ethical approval was granted by the Institutional Review Board (IRB) of the Harvard Faculty of Medicine at Harvard Medical School in the USA. Written informed consent was obtained from adolescents 18 years of age and over or from guardian(s) of adolescents <18 years. Participants <18 years provided informed assent. Three adolescents <18 years without a guardian provided assent and were granted a waiver of consent.

## Results

We enrolled 30 ALWH (ten per hospital), with a median age of 19 years [interquartile range (IQR): 18-20]. The cohort included 19 (63%) ALWH who acquired HIV in early childhood, 13 (43%) ALWH who identified as female (including one transgender women), 10 (33%) men who reported having sex with other men, five (17%) Venezuelan nationals, and four young pregnant women (13%) (Table 1). Six (20%) had been lost from care at the time of enrollment (Table 1).

**Table 1.**
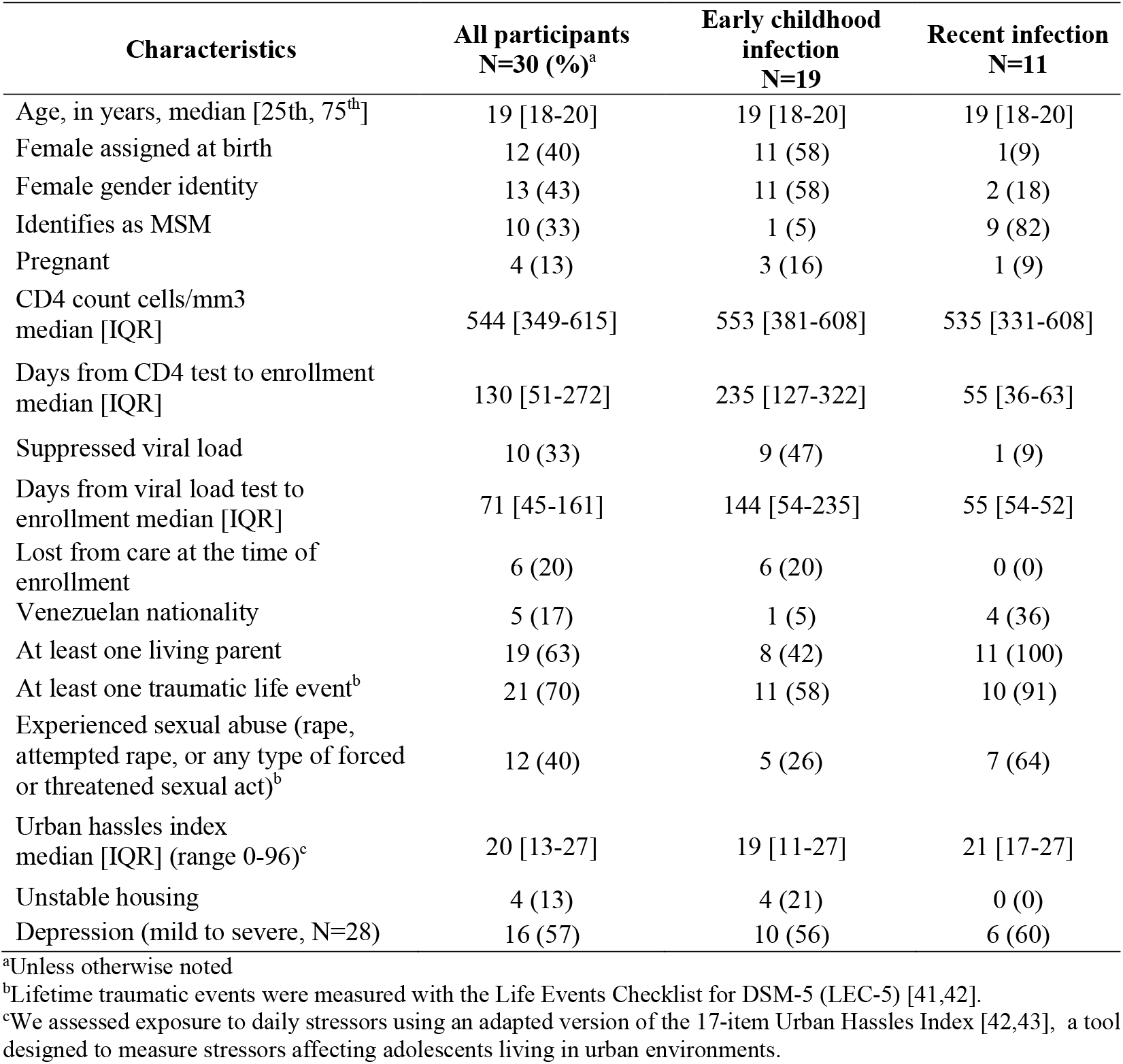
Baseline characteristics of adolescent participants transitioning to adult HIV care, Lima, Peru (N=30)^a^.

### Feasibility indicators

Of 36 adolescents referred for participation, six refused to participate (16.7%). Four were adolescents who had previously withdrawn from HIV care and did not wish to reengage. Two opted out because of scheduling conflicts with study activities. Once enrolled, no participant withdrew from the intervention, study, or ART. Of the 30 adolescents, all but three (90%) attended at least one in-person social support session (median among those participating in ≥1 session=2 [IQR: 2, 3] sessions). The median number of participants per in-person session was 7 [IQR: 6, 10]. There were 44 virtual sessions offered, and 28 adolescents (93%) participated in at least one of these sessions (median number of sessions attended by those who participated=34.5, [IQR:16.5-39.8]. The median number of participants per virtual session was 20 [IQR:18-21] and the median number of engagements during online sessions was 46 [IQR: 38, 59].

### Preliminary evidence of effectiveness

Changes in adherence scores, psychosocial scale scores, and transition readiness are shown in Table 2, Figure 3, Figure 4, Figure 5, and Appendices I-III. In transition readiness, we observed within-person improvements related to personal health (+1.9 points, p<0.001), healthcare usage (+2.4 points, p<0.001), knowledge (+3.3 points, p=0.001), and behavior (+3 points, p=0.003) at the end of the intervention, relative to baseline (Table 2). Results for adherence psychosocial constructs followed a pattern of modest improvements in each scale between baseline and six-months, which increased further by nine-months and were generally sustained at twelve months (Table 2). Although sample sizes were small for subgroup analyses, we observed evidence of improvements in transition readiness across all subgroups as well as improvements in adherence among adolescents living with HIV since early childhood, improvements in psychosocial indicators among adolescents with newly acquired HIV, and improvements in both metrics among adolescents with a history of non-adherence (Appendices I-III).

**Table 2.** Within-person changes in key outcomes among PASEO intervention participants (N=30)

| Outcome (range of possible values) | Score<br>median [IQR]<sup>a</sup> |  |  |  | Within- person change from baseline<br>median [IQR]<sup>a</sup> |  |  | P-value,<br>within- person change<sup>a</sup> |  |  |
| --- | --- | --- | --- | --- | --- | --- | --- | --- | --- | --- |
|  | Baseline | 6 mos. | 9 mos. | 12 mos. | 6 mos. | 9 mos. | 12 mos. | 6 mos. | 9 mos. | 12 mos. |
| **Self-reported adherence to ART** |  |  |  |  |  |  |  |  |  |  |
| Doses missed last 30 days, (0-30)<sup>b,c</sup> | 2 [0, 13] | 0.5 [0, 2] | 1 [0, 3] | 0.5 [0, 2] | 0 [-11, 0] | -1 [-11, 0] | -1 [-12, 0] | 0.03<sup>d</sup> | 0.03<sup>d</sup> | 0.01<sup>d</sup> |
| How often did you take your ART medications correctly, last 30 days? (1-5)<sup>c,e</sup> | 4 [2, 5] | 5 [4, 5] | 4 [4, 5] | 5 [4, 5] | 1 [0, 2] | 0 [0, 2] | 1 [0, 2] | 0.01<sup>d</sup> | 0.01<sup>d</sup> | 0.02<sup>f</sup> |
| How do you consider that you took your ART medications, as directed by your doctor, last 30 days? (1-6)<sup>c,e</sup> | 4 [3, 4] | 5 [4, 6] | 4 [3, 5] | 5 [4, 5] | 1 [0, 2]<sup>c</sup> | 0 [-1, 2.] | 1 [0, 2] | 0.001<sup>d</sup> | 0.09<sup>f</sup> | 0.02<sup>f</sup> |
| **Psychosocial outcomes** |  |  |  |  |  |  |  |  |  |  |
| Emotional support (7-33)<sup>e</sup> | 20 [13, 29] | 22 [17, 31] | 24.8 [16, 31] | 25.8 [18.7, 32.7] | 1[-2, 5] | 1 [-2, 5] | 2 [-1, 10] | 0.21<sup>f</sup> | 0.10<sup>d</sup> | 0.03<sup>f</sup> |
| Instrumental support (8-40)<sup>e</sup> | 24 [16, 31] | 26 [19, 31] | 30 [21, 36]<sup>g</sup> | 27.5 [24, 34] | 0 [-3, 4.3] | 2.6 [-1, 6]<sup>g</sup> | 2 [0, 8] | 0.67<sup>d</sup> | 0.07<sup>d,g</sup> | 0.004<sup>d</sup> |
| Self-efficacy (10-40)<sup>e</sup> | 23 [19, 28] | 25.5 [20, 30] | 27 [21, 33]<sup>g</sup> | 24.7 [21, 31] | 1 [-4, 5] | 3 [-2, 6]<sup>g</sup> | 1.5 [-2, 8] | 0.57<sup>d</sup> | 0.03<sup>f,g</sup> | 0.06<sup>f</sup> |
| Perceived stress (10-40)<sup>b</sup> | 19 [17, 23] | 19.2 [18, 22] | 18 [15, 20] | 18 [17, 20] | -0.5 [-3, 3] | -1.5 [-4, 2] | -1 [-5, 2] | 0.59<sup>f</sup> | 0.09<sup>d</sup> | 0.04<sup>f</sup> |
| **Transition readiness<sup>h</sup>** |  |  |  |  |  |  |  |  |  |  |
| Got transition, my health (0-18)<sup>e</sup> | 13.8 [12, 16] | -- | 16 [15, 18] | -- | -- | 1.9 [0, 4] | -- | -- | <0.001<sup>f</sup> | -- |
| Got transition, health care usage (0-28)<sup>e</sup> | 21 [18, 23.7] | -- | 24.1 [22, 27] | -- | -- | 2.4 [1, 6] | -- | -- | <0.001<sup>f</sup> | -- |
| Am I ON TRAC, knowledge (12-60)<sup>e</sup> | 48 [38.7, 52] | -- | 50.5 [47, 55] | -- | -- | 3.3 [1, 8] | -- | -- | 0.001<sup>d</sup> | -- |
| Am I ON TRAC, behavior (9-45)<sup>e</sup> | 28 [24, 31] | -- | 32 [27, 35] | -- | -- | 3 [0, 6] | -- | -- | 0.003<sup>f</sup> | -- |

<sup>a</sup>6, 9, and 12 months after enrollment correspond to the end of the intensive phase, the end of the taper phase, and three months after the intervention, respectively
<sup>b</sup>Lower=favorable
<sup>c</sup>N=27 for baseline measurement and all within-person change comparisons. Three participants on ART for less than 7 days at the time of baseline data collection were excluded.
<sup>d</sup>Wilcoxon signed rank
<sup>e</sup>Higher=favorable
<sup>f</sup> Paired T-test
<sup>g</sup>N=29
<sup>h</sup>Transition readiness was assessed at baseline and 9 months

**Figure 3.**
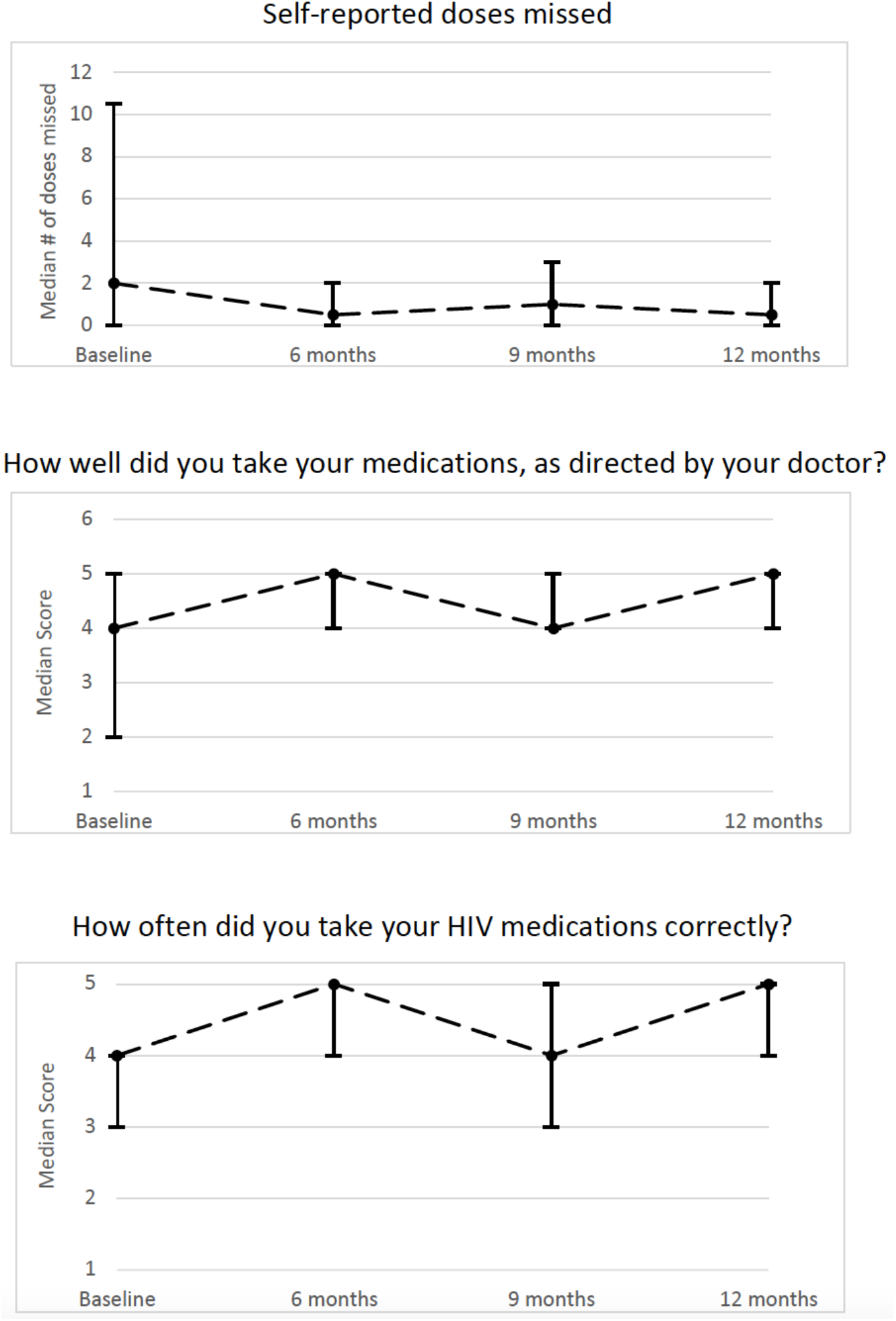
Treatment adherence in the last 30 days among adolescents living with HIV in Lima, Peru. Circles indicate the mean; error bars indicate the interquartile range

**Figure 4.**
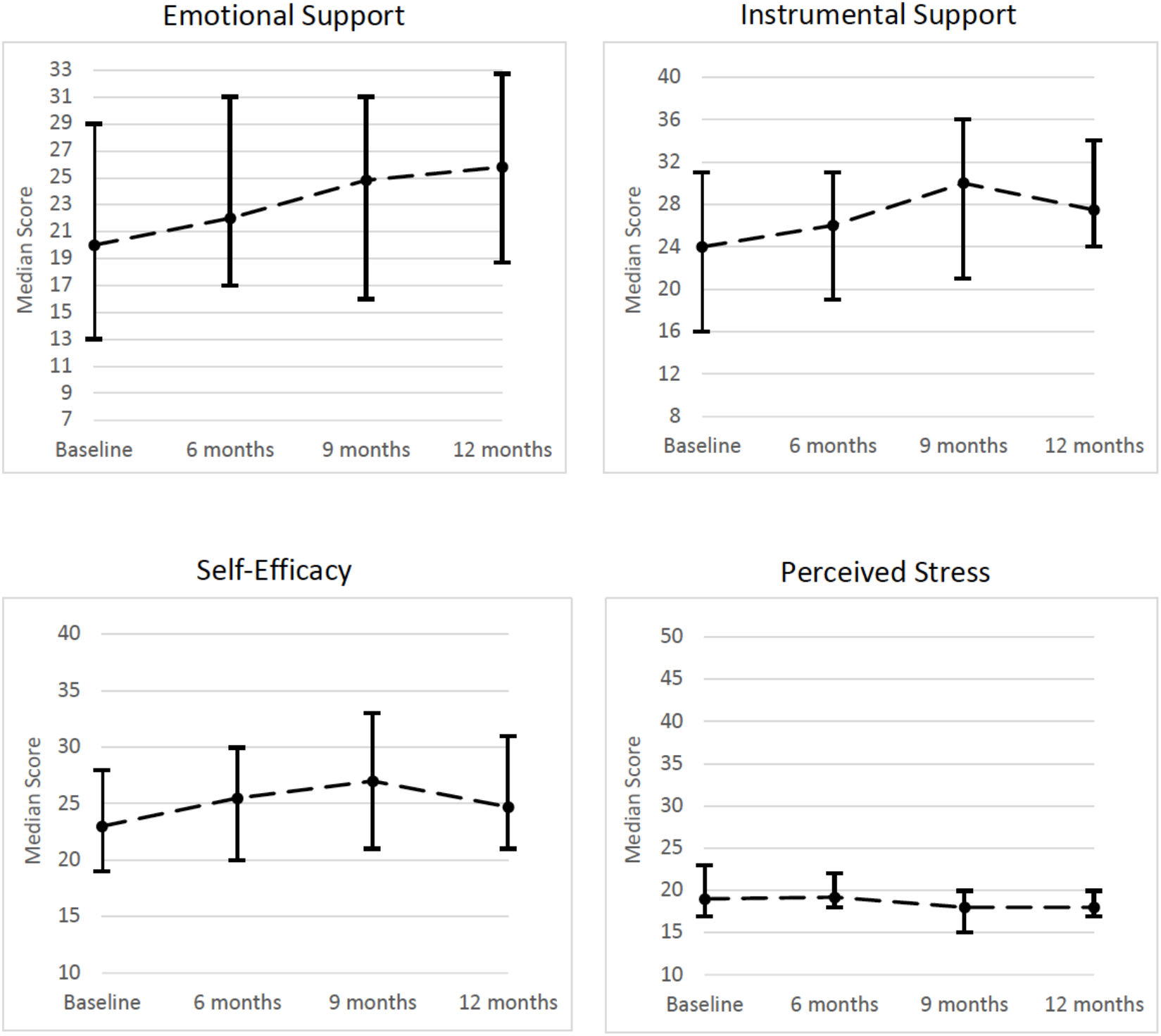
Psychosocial outcome scores among adolescents living with HIV in Lima, Peru. Circles indicate the mean; error bars indicate the interquartile range

**Figure 5.**
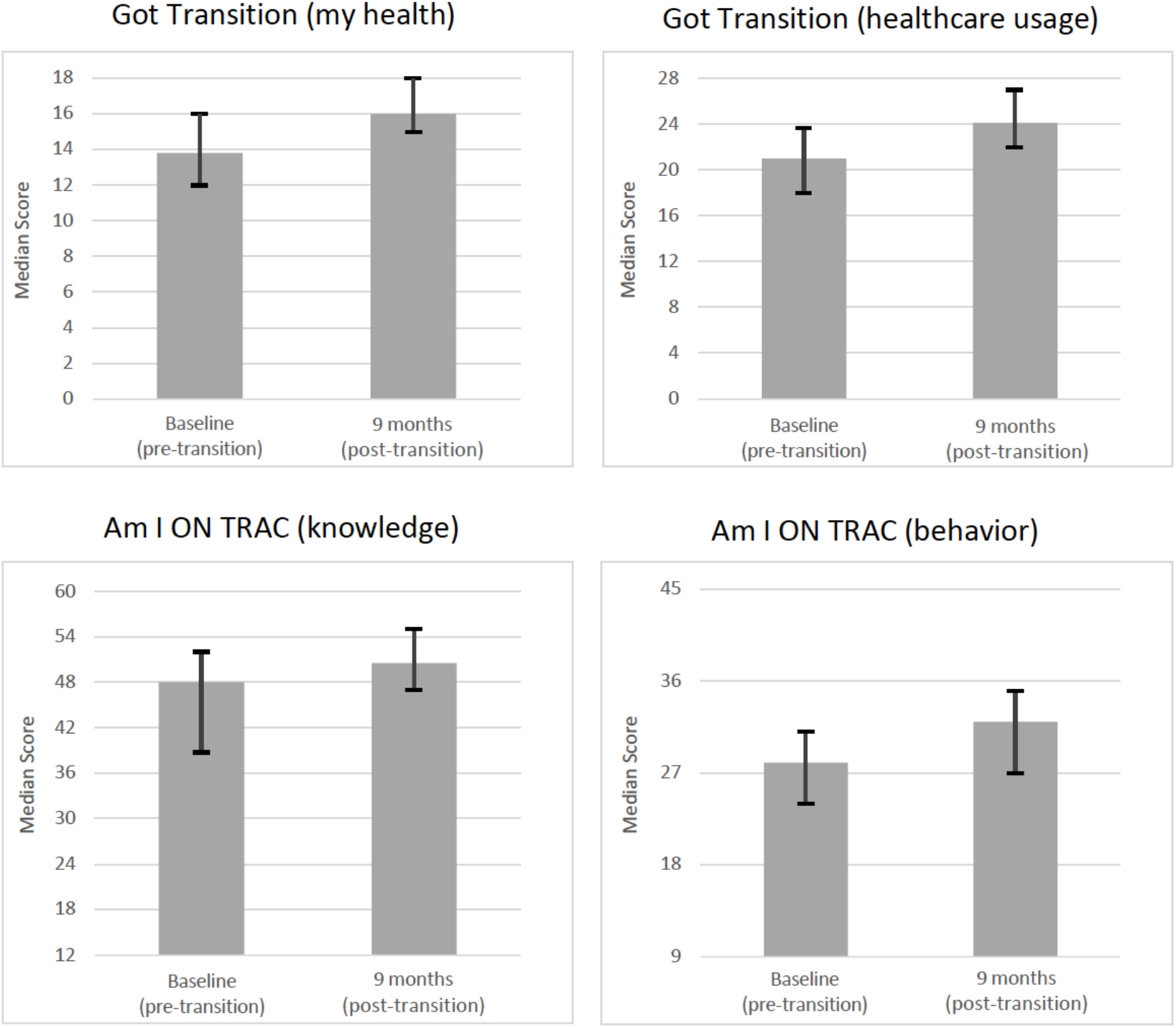
Changes in transition readiness among adolescents living with HIV in Lima, Peru. The error bars indicate the interquartile range

## Discussion

We provide quantitative evidence supporting the feasibility and effectiveness of a community-based intervention designed to improve health outcomes for ALWH in Lima, Peru. The adolescents in this cohort were diverse in terms of their medical needs and clinical circumstances, which included depression, housing instability, poverty, lack of familial support, pregnancy and living in a foreign country. A key strength of the intervention was its comprehensive nature (i.e., it was designed to address a broad spectrum of unmet social and health needs) and individualized design (i.e., it sought to respond to each adolescent’s needs, tailoring the intensity of support offered to each participant). Research has highlighted the limited efficacy of a “one size fits all” approach for community-based interventions [8,44,45], in part because subgroups at highest risk may need extra support [46,47]. In this intervention, entry-level health workers filled gaps in care, provided linkages, liaised with providers, and literally accompanied adolescents as they transitioned to adult care.

We found the PASEO intervention to be highly feasible based on metrics of acceptability and demand. There was a low refusal rate for enrollment, no participants were lost to follow up, and adolescents demonstrated high-attendance and engagement in group sessions. The potential effectiveness of PASEO is supported by within-person improvements in adherence, psychosocial indicators, and transition readiness; this improvement was often sustained three months after the intervention’s intensive phase. Importantly, these improvements appear to hold up across subgroups of adolescents characterized by timing of diagnosis (early childhood infection versus recent diagnosis) and among those with a history of non-adherence. This suggests that the lay health workers were effective at individualizing support to adequately meet the needs of each adolescent. Importantly, these data also corroborate that PASEO aligns with World Health Organization criteria for adolescent-friendly care in that it was acceptable (adolescents were willing to engage), equitable (all adolescents could access it), appropriate (offering the right menu of services) and effective (resulting in a positive contribution to health) [48]. To further increase equity, adaptations may be needed to ensure it is acceptable and effective for transgender women, as we only enrolled one in this study.

A number of intervention studies addressing the transition period are underway in low-or-middle-income and high-burden settings [49,50]. While these studies will contribute importantly to knowledge on evidence-based practices, there is a notable lack of intervention studies addressing the transition to adult care in South America. This disparity reflects a broader knowledge gap related to effective interventions for ALWH in the Latin American and Caribbean contexts more generally. One recent systematic review of interventions for ALWH found only six studies that discussed linkage to care, none of which were from Latin America [51]; another, focused on psycho-social support interventions, identified six studies, but found none outside the U.S. or Africa. An intervention that was effective in improving continuity of care and psychosocial health outcomes among ALWH in South America would therefore contribute to filling an important research and implementation gap. And, the fact that the six adolescents who had been lost from care at enrollment remained engaged in ART at the end of the study, suggests that the PASEO intervention also may be an effective strategy for re-engaging ALWH who are lost from care.

This pilot study was underway when the SARS-CoV-2 pandemic emerged, during which stay-at-home orders were intermittently instated. These circumstances required us to adapt the intervention to a virtual format; however, improvements in key metrics suggests that the pillars of the intervention (accompaniment, social support, skills-building) were preserved under this hybrid model. The lack of a control group precludes definitively concluding that the intervention caused these improvements; however, baseline measurements preceded SARS-CoV-2, while follow-up measurements occurred in the first six months of the pandemic; therefore, the expected underlying time trend in these outcomes would be downward and would likely have attenuated any PASEO-related improvements [31]. While unplanned, the intervention adaptations could be considered a strength: the flexibility and high participation rates despite the pandemic suggest that the feasibility metrics are even more notable. And, successful adaptations open the possibility for flexible delivery across a variety of in-person parameters. The pandemic also resulted in temporary suspension of CD4 cell count and viral load testing, which prevented us assessing biologic endpoints.

## Conclusions

In conclusion, the PASEO intervention was feasible and demonstrated promise for improving myriad self-reported health outcomes in a diverse cohort of ALWH. Future studies should include a large-scale impact evaluation including biological outcomes and assessment of long-term effectiveness beyond the life of the intervention.

## Supporting information

Research reporting checklist

Beckhorn author disclosure

Errea author disclosure

ESanchez author disclosure

Galea author disclosure

HSanchez author disclosure

Kolevic author disclosure

KRamos author disclosure

Lecca author disclosure

Lindeborg author disclosure

Matos author disclosure

Rodriguez author disclosure

Senador author disclosure

Shin author disclosure

Vargas author disclosure

Wong author disclosure

Franke author disclosure

## Data Availability

The authors confirm that the data supporting the findings of this study are available within the article and its supplementary material.

## Competing interests

The authors declare no competing interests.

## Authors’ contributions

Conceived of the study: MF

Developed the study intervention: MW, SS, HS, LL, MF, JG

Designed the study: MF, MW, JG, SS

Oversaw enrollment and recruitment: MW, LK, EM, ES

Implemented the intervention: MW, HS, LS, AR, RE, KR

Data collection and management: LS, AR, RE, MW, CR, AL, VV, CB

Analyzed data: VV, CR, AL

Interpreted data: all authors

Wrote first draft of the manuscript: VV, MF

Critically reviewed and approve the final version of the manuscript: all authors

## Acknowledgements

We acknowledge and thank the adolescents who participated in the study; personnel at Hospital Nacional Hipólito Unanue, Hospital Nacional Arzobispo Loayza, and the Instituto Nacional de Salud del Niño and the Peru National HIV Program. We are indebted to the Socios En Salud Youth Advisory Board for their feedback and contributions.

## Funding

This research was entirely supported by the National Institute of Allergy and Infectious Diseases of the National Institutes of Health under award number R21AI143365.

## Disclaimer

The content is solely the responsibility of the authors and does not necessarily represent the official views of the National Institutes of Health.

## Additional files

Additional file 1: Appendix

Information on file format. Analyses of effectiveness outcomes, stratified by early childhood versus recent diagnosis and whether the adolescent had been lost from care or had a history of chronic non-adherence.

## List of abbreviations

ALWH: adolescents living with HIV,
ART: antiretroviral therapy,
CBA: community-based accompaniment,
CHWs: community health workers,
DOT: directly observed treatment,
HIV: human immunodeficiency virus,
MSM: men who have sex with men,
PLWH: people living with HIV,
SES: Socios En Salud,
SMS: short message service,
TGW: transgender women,
YAB: youth advisory board

## Notes

### Competing Interest Statement

The authors have declared no competing interest.

### Clinical Trial

NCT05022706

### Author Declarations

Ethical approval was granted by the Institutional Research Ethics Committees of the Instituto Nacional de Salud del Nino, Hospital Nacional Arzobispo Loayza, and Hospital Nacional Hipolito Unanue in Peru. In addition, ethical approval was granted by the Institutional Review Board (IRB) of the Harvard Faculty of Medicine at Harvard Medical School in the USA.

