## Appendix for "Community-Based Accompaniment for Adolescents Transitioning to Adult HIV Care in Urban Peru: a Pilot Study"

### Adaptation of the "Am I on TRAC" transition readiness questionnaire.

The 21-item questionnaire consists of a knowledge index and a behavior index. The original knowledge subscale consisted of 16 questions and four response categories, ranging from "strongly disagree" to "strongly agree". However, based on feedback from the Youth Advisory Board, we added a "neutral" response item and removed four questions that were less applicable to the study population ("I know how to get my medical records", "I know how my health condition might limit my career choices", "I know how my health condition affects my physical activities", and "I have a family doctor").

**Appendix I. Changes in key outcomes among PASEO intervention participants who were lost from care or had a history of chronic non-adherence to ART (N=13)**

| Outcome (range of possible values) | Score median [IQR][a] | | | | Within- person change from baseline median [IQR][a] | | | P-value, within- person change[a] | | |
|---|---|---|---|---|---|---|---|---|---|---|
| | Baseline | 6 mos. | 9 mos. | 12 mos. | 6 mos. | 9 mos. | 12 mos. | 6 mos. | 9 mos. | 12 mos. |
| **Self-reported adherence to ART** | | | | | | | | | | |
| Doses missed last 30 days, (0-30)[b] | 5 [2, 30] | 0 [0, 2] | 1 [0, 3] | 0 [0, 2] | -5 [-29, 0] | -5 [-29, 0] | -3 [-30, -1] | 0.02[c] | 0.02[c] | 0.002[c] |
| How often did you take your ART medications correctly? last 30 days, (1-5)[d] | 3 [2, 4] | 5 [4, 5] | 4 [3, 5] | 5 [3, 5] | 2 [0, 3] | 1 [0, 3] | 1 [0, 2] | 0.02[c] | 0.04[c] | 0.04[c] |
| How do you consider that you took your ART medications, as directed by your doctor? last 30 days (1-6)[d] | 3 [2, 4] | 4 [4, 5] | 5 [3, 5] | 5 [4, 5] | 1 [03] | 1 [0, 3] | 1 [0, 2] | 0.02[e] | 0.09[e] | 0.008[c] |
| **Psychosocial outcomes** | | | | | | | | | | |
| Emotional support (7-33)[d] | 21 [13, 26] | 17 [12,28] | 18.7 [14,27] | 19 [18, 33] | 0 [-3, 5] | 1 [-2.3, 3] | 5 [-2.3, 8] | 0.84[e] | 0.81[e] | 0.24[e] |
| Instrumental support (8-40)[d] | 20 [15,26] | 20 [16,22] | 27.8 [16.5, 31] | 24 [20, 27] | 0 [-3, 4.3] | 3.5 [-1, 4.5] | 4 [2, 7] | 0.81[c] | 0.09[e] | 0.002[e] |
| Self-efficacy (10-40)[d] | 20 [19,24] | 21 [16,28] | 24 [21,30.5] | 21.1 [19, 26] | 1 [-3.7, 7] | 4 [-2.5, 10.5][f] | 2 [-2, 7] | 0.33[e] | 0.07[e,f] | 0.18[e] |
| Perceived stress (10-40)[b] | 20 [18,23] | 19.2 [18,22] | 19 [17,20] | 19 [17, 20] | 0 [-3, 3] | -1 [-2, 2] | -0.7 [-5, 2] | 0.82[e] | 0.38[e] | 0.17[e] |
| **Transition readiness** | | | | | | | | | | |
| Got transition, my health 0-18)[d,g] | 14 [12,15] | - | 15.7 [15,18] | - | - | 1.2 [0.6, 3] | - | - | 0.01[e] | - |
| Got transition, health care usage (0-28)[d,g] | 22 [21,24] | - | 24.1 [23,28] | - | - | 2 [1, 5.1] | - | - | 0.001[c] | - |
| Am I ON TRAC, knowledge (12-60)[d,g] | 51 [46,52] | - | 54 [48,58] | - | - | 4 [2, 6.6] | - | - | <0.001[e] | - |
| Am I ON TRAC, behavior (9-45)[d,g] | 28 [24,29] | - | 28 [26,32] | - | - | 0 [-2, 6] | - | - | 0.29[e] | - |

[a] 6, 9, and 12 months after enrollment correspond to the end of the intensive phase, the end of the taper phase, and three months after the intervention, respectively
[b] Lower=favorable
[c] Wilcoxon signed rank
[d] Higher=favorable
[e] Paired T-test
[f] N=12
[g] Transition readiness was assessed at baseline and 9 months

**Appendix II. Changes in key outcomes among PASEO intervention participants with recent HIV diagnosis (N=11)**

| Outcome (range of possible values) | Score median [IQR][a] | | | | Within- person change from baseline median [IQR] [a] | | | P-value, within- person change [a] | | |
|---|---|---|---|---|---|---|---|---|---|---|
| | Baseline | 6 mos. | 9 mos. | 12 mos. | 6 mos. | 9 mos. | 12 mos. | 6 mos. | 9 mos. | 12 mos. |
| **Self-reported adherence to ART** | | | | | | | | | | |
| Doses missed last 30 days, (0-30)[b,c] | 0.5 [0, 5] | 1.5 [0, 3] | 1 [0, 2.5] | 1 [1, 3.5] | 0 [-4, 3] | 0 [-5, 2] | 0 [-3.5, 3] | 0.83[d] | 1[f] | 1[f] |
| How often did you take your ART medications correctly? last 30 days, (1-5)[e,c] | 5 [4, 5] | 5 [4, 5] | 4 [4, 5] | 5 [4, 5] | 0.5 [-0.5, 1] | 0 [-0.5, 0.5] | 0.5 [-1, 1] | 0.14[d] | 0.80[d] | 0.84[d] |
| How do you consider that you took your ART medications, as directed by your doctor? last 30 days (1-6) [e,c] | 4.5 [4, 5] | 5 [4.5, 5] | 4.5 [4, 5] | 5 [4, 5] | 0.5 [0, 1.5] | 0 [-0.5, 1] | 0.5 [-1, 1] | 0.35[d] | 1[d] | 0.82[d] |
| **Psychosocial outcomes** | | | | | | | | | | |
| Emotional support (7-33) [e] | 18 [11, 30] | 26 [22, 31] | 29 [21, 31] | 31 [23, 32.7] | 1 [0, 13] | 2 [0, 18] | 3 [0.7, 20] | 0.05[f] | 0.02[f] | 0.04[f] |
| Instrumental support (8-40) [e] | 24 [16, 31] | 29 [25, 34] | 30 [25, 37] | 32 [27, 37] | 2 [-2, 12] | 5 [-1, 10] | 5 [2, 16] | 0.11[d] | 0.11[d] | 0.02[d] |
| Self-efficacy (10-40) [e] | 25 [19, 32] | 26 [24, 29] | 28 [20, 37] | 30 [24.4, 36] | 2 [-5, 5] | 2 [-2, 4] | 2 [-2, 14] | 0.45[d] | 0.28[f] | 0.05[d] |
| Perceived stress (10-40) [e] | 19 [17, 25] | 20 [17, 22] | 20 [15, 22] | 18 [18, 18] | -1 [-3, 3] | -1 [-4, 2] | -2 [-7, -1] | 0.46[d] | 0.23[d] | 0.02[d] |
| **Transition readiness** | | | | | | | | | | |
| Got transition, my health 0-18) [e,g] | 12 [10, 16] | - | 15 [14, 16] | - | - | 2.3 [-1, 6] | - | - | 0.09[d] | - |
| Got transition, health care usage (0-28)[e,g] | 19 [17, 23] | - | 23 [19, 27] | - | - | 2 [-0.5, 6] | - | - | 0.06[d] | - |
| Am I ON TRAC, knowledge (12-60) [e,g] | 43 [38, 49.1] | - | 50 [42, 52] | - | - | 3 [0, 8] | - | - | 0.07[d] | - |
| Am I ON TRAC, behavior (9-45) [e,g] | 28 [23, 29] | - | 34 [32, 38] | - | - | 6 [3, 9] | - | - | 0.008[d] | - |

[a] 6, 9, and 12 months after enrollment correspond to the end of the intensive phase, the end of the taper phase, and three months after the intervention, respectively
[b] Lower=favorable
[c] N=8 participants who had been on cART for at least 30 days at baseline
[d] Paired T-test
[e] Higher=favorable
[f] Wilcoxon signed rank
[g] Transition readiness was assessed at baseline and 9 months

**Appendix III. Changes in key outcomes among PASEO intervention participants with early childhood HIV infection (N=19)**

| Outcome (range of possible values) | Score median [IQR][a] | | | | Within- person change from baseline median [IQR] [a] | | | P-value, within- person change [a] | | |
|---|---|---|---|---|---|---|---|---|---|---|
| | Baseline | 6 mos. | 9 mos. | 12 mos. | 6 mos. | 9 mos. | 12 mos. | 6 mos. | 9 mos. | 12 mos. |
| **Self-reported adherence to ART** | | | | | | | | | | |
| Doses missed last 30 days, (0-30) [b] | 3 [0, 30] | 0 [0, 2] | 1 [0, 3] | 0 [0, 2] | -1 [-28, 0] | -1 [-17, 0] | -1 [-25, 0] | 0.02[c] | 0.01[c] | <0.001[c] |
| How often did you take your ART medications correctly? last 30 days, (1-5) [d] | 3 [2, 5] | 5 [4, 5] | 4 [4, 5] | 4 [3, 5] | 1 [0, 3] | 1 [0, 3] | 1 [0, 3] | 0.03[c] | 0.006[c] | 0.02[c] |
| How do you consider that you took your ART medications, as directed by your doctor? last 30 days (1-6) [d] | 3 [2, 4] | 5 [4, 6] | 5 [3, 5] | 5 [4, 5] | 1 [0, 3] | 1 [-1, 3] | 1 [0, 2] | 0.004[e] | 0.09[e] | 0.002[c] |
| **Psychosocial outcomes** | | | | | | | | | | |
| Emotional support (7-33) [d] | 21 [13, 29] | 19 [13, 31] | 19 [14, 31] | 19 [17, 33] | 0 [-4, 5] | 0 [-2.3, 3] | 1 [-2.3, 8] | 0.99[e] | 0.66[e] | 0.24[e] |
| Instrumental support (8-40) [d] | 23 [15, 36] | 21 [17.3, 31] | 29.3 [19, 35][f] | 24 [20, 33] | 0 [-4, 1] | 1.8 [-2, 4][f] | 2 [-1.9, 7] | 0.47[e] | 0.29[e,f] | 0.21[e] |
| Self-efficacy (10-40) [d] | 23 [19, 26] | 22 [16, 31] | 24.5 [21, 31][f] | 23 [19, 28] | -1 [-3.7, 6] | 3 [-2, 9] | 0 [-2, 8] | 0.46[e] | 0.09[e] | 0.35[e] |
| Perceived stress (10-40) [b] | 19 [16, 23] | 19 [18, 22] | 17 [15, 20] | 18 [16, 21] | 0 [-4, 3] | -2 [-4, 2] | -0.7 [-5, 2] | 0.74[e] | 0.12[e] | 0.20[e] |
| **Transition readiness** | | | | | | | | | | |
| Got transition, my health 0-18) [d,g] | 14 [12.3, 16.8] | - | 17 [15, 18] | - | - | 1.7 [0.6, 3.7] | - | - | 0.001[e] | - |
| Got transition, health care usage (0-28) [d,g] | 22 [19, 24] | - | 25 [23, 28] | - | - | 2.7 [1, 6] | - | - | <0.001[e] | - |
| Am I ON TRAC, knowledge (12-60) [d,g] | 51 [46, 54] | - | 51 [48, 59] | - | - | 3.5 [1, 7] | - | - | 0.006[c] | - |
| Am I ON TRAC, behavior (9-45)[d,g] | 28 [24, 32] | - | 30 [26, 35] | - | - | 0 [-2, 6] | - | - | 0.16[e] | - |

[a] 6, 9, and 12 months after enrollment correspond to the end of the intensive phase, the end of the taper phase, and three months after the intervention, respectively
[b] Lower=favorable
[c] Wilcoxon signed rank
[d] Higher=favorable
[e] Paired T-test
[f] N=18
[g] Transition readiness was assessed at baseline and 9 months
